# Prevalence of SARS-CoV-2 infection among people experiencing homelessness in Toronto during the first wave of the COVID-19 pandemic

**DOI:** 10.1101/2021.09.21.21263713

**Authors:** Linh Luong, Michaela Beder, Rosane Nisenbaum, Aaron Orkin, Jonathan Wong, Cynthia Damba, Ryan Emond, Suvendrini Lena, Vanessa Wright, Mona Loutfy, Cindy Bruce-Barrett, Wilfred Cheung, Yick Kan Cheung, Victoria Williams, Miriam Vanmeurs, Andrew Boozary, Harvey Manning, Joe Hester, Stephen W. Hwang

**Author notes:** **Declarations:**. **Ethics approval:** This is a research study without requiring consent from individuals to whom the information relates. No individual-level information or personal health information were collected and the original data is in aggregate format. All procedures performed in studies involving human participants were in accordance with the ethical standards of the institutional and/or national research committee (Research Ethics Board of St. Michael’s Hospital, Unity Health Toronto, in Toronto, Ontario, Canada. REB#20-158) and with the 1964 Helsinki declaration and its later amendments or comparable ethical standards. **Author Contributions:** Stephen Hwang and Michaela Beder contributed to the creation of the study and its design. Rosane Nisenbaum and Linh Luong performed the statistical analysis. Rosane Nisenbaum, Linh Luong, and Stephen Hwang interpreted the results. The initial draft of the manuscript was written by Linh Luong and Stephen Hwang. All authors contributed to the revisions of the manuscript and approved the final version. **Availability of data and material:** Not applicable. **Consent to participate:** Not applicable. **Consent to publication:** Not applicable. **Code availability:** Not applicable.

## Abstract

**Background:** People experiencing homelessness are at increased risk of SARS-CoV-2 infection. This study reports the point prevalence of SARS-CoV-2 infection during testing conducted at sites serving people experiencing homelessness in Toronto during the first wave of the COVID-19 pandemic. We also explored the association between site characteristics and prevalence rates.

**Methods:** The study included individuals who were staying at shelters, encampments, COVID-19 physical distancing sites, and drop-in and respite sites and completed outreach-based testing for SARS-CoV-2 during the period April 17 to July 31, 2020. We examined test positivity rates over time and compared them to rates in the general population of Toronto. Negative binomial regression was used to examine the relationship between each shelter-level characteristic and SARS-CoV-2 positivity rates. We also compared the rates across 3 time periods (T1: April 17-April 25; T2: April 26-May 23; T3: May 24-June 25).

**Results:** The overall prevalence of SARS-CoV-2 infection was 8.5% (394/4657). Site-specific rates showed great heterogeneity with infection rates ranging from 0% to 70.6%. Compared to T1, positivity rates were 0.21 times lower (95% CI: 0.06, 0.75) during T2 and 0.14 times lower (95% CI: 0.043, 0.44) during T3. Most cases were detected during outbreak testing (384/394 [97.5%]) rather than active case finding.

**Interpretation:** During the first wave of the pandemic, rates of SARS-CoV-2 infection at sites for people experiencing homelessness in Toronto varied significantly over time. The observation of lower rates at certain sites may be attributable to overall time trends, expansion of outreach-based testing to include sites without known outbreaks and/or individual site characteristics.

## Objectives

The COVID-19 pandemic has disproportionately impacted and continued to pose extraordinary challenges for people experiencing homelessness (Perri et al., 2020; Turnbull et al., 2021), especially those from Indigenous and racialized communities. Prior to the pandemic, many of these individuals were already struggling due to low social assistance rates, the opiate crisis, and a chronic lack of sufficient shelter beds. More than 35,000 people experience homelessness on any given night in Canada, and at least 235,000 individuals were homeless in a year (Gaetz et al., 2016). Indigenous and racial/ethnic minority groups are overrepresented in the homeless population, constituting about 10% and 54%, respectively, of homeless population in Toronto (City of Toronto, 2018). People experiencing homelessness are at increased risk for infectious diseases because of their poor health and complex health concerns ranging from physical to mental illnesses and addiction and substance-use issues (Hwang, 2001). In addition, crowding and shared living spaces in shelters make it difficult for residents to adhere to pandemic-related physical distancing guidelines. Further, the number of homeless persons living in outdoor encampments has risen during the pandemic, raising questions about whether the risk for SARS-CoV-2 infection in encampments is higher or lower than in shelters. Few studies have examined SARS-CoV-2 infection in people experiencing homelessness in both shelters and encampments in Canada.

The objective of this study was to estimate the prevalence of SARS-CoV-2 infection among people experiencing homelessness in the City of Toronto who received outreach-based testing at shelters, drop-ins and encampments during the first COVID-19 pandemic wave. We also explored the association between site characteristics and prevalence rates.

## Methods

### Study design and setting

This study is a retrospective review of aggregated data collected by Ontario Health Toronto as part of outreach testing for SARS-CoV-2 infection among individuals experiencing homelessness in Toronto. This was a combination of two testing programs: 1) an active case finding program involving testing in shelters without identified cases throughout the study period, and 2) an outbreak management program involving testing in response to positive cases that were identified through community testing or symptoms. Testing was conducted across various types of sites, including shelter-based programs, drop-in and respite programs, encampments and COVID-19 physical distancing sites, which are sites that were opened during the pandemic to allow safe physical distancing for shelter residents. The study included data on testing performed on residents at these sites between April 17 and July 31, 2020, which included the first wave of Ontario’s COVID-19 pandemic. Each on-site testing was done either for active case finding or for outbreak management purpose. Outbreak was defined as having at least one laboratory confirmed case in residents or staff. This is in accordance with the Ontario Ministry of Health’s outbreak guidance for congregate settings(Ministry of Health and Long-Term Care, 2020). Sites not indicated for outbreak testing were classified as active case finding.

### Participant

Shelters, drop-ins and respite sites, encampments and COVID-19 physical distancing sites that completed nasopharyngeal swab tests for SARS-CoV-2 infection between April 17 and July 31, 2020 were eligible to be included in our study. We excluded testing at boarding homes, transitional housing, and permanent housing. We also excluded dedicated COVID-19 isolation and recovery sites as all individuals at these sites were either confirmed positive, presenting with COVID-19 symptomology, or were close contacts with a positive case prior to their admission to the sites. As many sites were tested more than once during the study, we only included results from the first testing period for each site to reduce double counting of individuals tested.

### Data sources

We used aggregated data from Ontario Health Toronto, Toronto Public Health (TPH), and the City of Toronto Shelter, Support and Housing Administration (SSHA) to ascertain outbreak status, COVID-19 test results and shelter characteristics. Ontario Health Toronto collected information on test dates, total number of residents tested, the number tested positive for each date, and the location of each site. TPH data were used to determine outbreak status for each testing, as well as weekly laboratory positivity rates (per 100 individuals tested) of the Toronto general population, and the number of sporadic cases by neighborhood regions (City of Toronto, 2020). The weekly positivity rate in the general population in Toronto was defined as the number of people who had a COVID-19 positive test result per 100 people tested each week (Sunday to Saturday) (City of Toronto, 2020). We collected shelter characteristics such as layout style and population type of each site using data from SSHA.

### Primary outcome

The primary outcome was the prevalence of SARS-CoV-2 infection at each site between April 17 and July 31, 2021. The prevalence was defined as the number of confirmed positive cases divided by total number of residents tested on each date. Sites with a large number of residents were tested over two or more days.

### Site Characteristics

Sites were grouped into four mutually exclusive categories: shelter, drop-in/respite, encampment, and COVID-19 physical distancing site. Shelter population was categorized by groups (youth, single adults and families) and gender (men, women and mixed). We further categorized site type as to whether or not it served exclusively refugee claimants. For each site, we defined its layout as one of the following: single room, shared room (2-5 beds), dorm or open style (more than 6 beds), and encampment (outdoor). If a site had a mixed layout (both single rooms and shared rooms), we classified the layout as shared rooms to reflect COVD-19 transmission risk in shared spaces. As a proxy measure of COVID-19 transmission in the community that could potentially affect site positivity rates, data were obtained on the number of sporadic cases of COVID-19 (i.e., cases not associated with outbreaks in healthcare and congregate settings) in the neighborhood where the sites were located.

### Statistical analyses

Prevalence of SARS-CoV-2 infection was expressed as positivity rates per 100 tested residents between April 17 and July 31, 2021, and it is described by site and by site characteristics. The overall prevalence of SARS-CoV-2 infection was calculated by dividing the total number of confirmed cases across all sites by the total number of tested individuals during the study period with 95% confidence interval. We also displayed the weekly COVID-19 positivity rates among individuals screened at sites for people experiencing homelessness in relationship to the positivity rates in the general Toronto population.

The association between site characteristics and site prevalence rates was evaluated using a negative binomial regression. The model’s outcome was the total number of positive cases in each site and included one site characteristic and the log of the total number of residents tested as the offset. Only univariate models were fit, since most the site characteristics were highly correlated. For our regression analysis, we focused on data between April 17 and June 25 since this period constituted most of the first wave of the pandemic. In addition, on June 25^th^, Toronto entered Stage 2 wherein businesses and services were allowed to reopen(Government of Ontario, 2020). We categorized our testing dates into 3 periods, T1: April 17-April 25; T2: April 26-May 23; T3: May 24-June 25. Rate ratios ratio (RR) and 95% confidence interval (CI) estimated ratios of daily prevalence rates. To compare outbreaks in homeless sites with sporadic cases in community, for each site, we calculated the cumulative sporadic cases from January 21 up until the date of the testing by Toronto neighborhood regions. All analyses were performed using SAS 9.4 (SAS Institute Inc. Cary NC, USA).

The study was approved by the Research Ethics Board of St. Michael’s Hospital, Unity Health Toronto, Canada.

## Results

The final sample included 97 unique sites and 111 testing dates. Eight sites had to perform testing on multiple days because of high resident volume which led to purposefully testing over more than 1 day, or because not all residents were available on the first day of testing. Among the excluded testing dates (n=49), 8 had missing test results, 23 were not at a site for people experiencing homelessness, and 18 were repeated testing.

Table 1 presents characteristics of the sites and testing completed between April 17 and July 31. The majority were shelter sites (58.8%). Most sites had a shared room layout (43.8%). At 92% of sites, testing was performed on only a single date. Testing was performed in response to an outbreak on 26.8% of the dates and for active case finding purposes (in the absence of any known outbreak) in 73.2% of the dates. In particular, between April 17 and June 25, there were 69 testing dates, and 58.0% of them were for outbreak purpose. After June 25, all 42 testing dates were performed for active case finding purposes.

**Table 1:** Testing characteristics of study sites between April 17 and July 31, 2020

| Description | N (%) |
| --- | --- |
| Number of Sites | 97 |
| Sector |  |
| Shelter | 57 (58.8) |
| Drop-in and respite programs | 12 (12.4) |
| Encampment site | 9 (9.3) |
| COVID-19 response sites and hotels | 19 (19.6) |
| Population group |  |
| Youth | 8 (8.3) |
| Single adults | 79 (81.4) |
| Families | 10 (10.3) |
| Population gender |  |
| Men | 25 (25.8) |
| Women | 22 (22.7) |
| Mixed | 50 (51.6) |
| Refugee sites | 7 (7.2) |
| Reason for testing |  |
| Outbreak | 26 (26.8) |
| Active case finding | 71 (73.2) |
| Layout<sup>†</sup> |  |
| Single room | 20 (20.8) |
| Shared room | 42 (43.8) |
| Dorm style or open layout | 25 (26.0) |
| Encampment | 9 (9.4) |
| Number of testing conducted by time period (n=111) |  |
| 17 Apr - 25 Apr | 8 (7.2) |
| 26 Apr - 23 May | 19 (17.1) |
| 24 May - 20 Jun | 31 (27.9) |
| 21 Jun - 18 Jul | 42 (37.8) |
| 19 Jul - 31 Jul | 11 (9.91) |

<sup>†</sup>Missing for one site

The overall prevalence of SARS-CoV-2 infection among shelter residents between April 17 and July 31 was 8.5% (394/4657, 95% CI 7.7% - 9.3%). For the period of April 17 to June 25, the prevalence was 11.5% (391/3415, 95% CI 10.4% - 12.5%). Site-specific rates ranged from 0% to 70.6% and declined over time (Figure 1). After June 20, the prevalence was essentially zero. Most cases were detected during outbreak testing (384/394 [97.5%]) rather than active case finding. Compared to the City of Toronto epidemic curve, the Toronto shelter positivity rates were consistently higher, with the exception of the period between April 26 and May 10 (Figure 2). All youth shelters were tested after May 24, 2020, and their SARS-CoV-2 infection rates were essentially zero. Given little variability in the youth shelters sample for comparison, we combined them with family shelters in our regression analysis. The daily prevalence rates of SARS-CoV-2 during T2 and T3 were significantly lower compared to the rates during T1 (Table 2). Compared to T1, T2 and T3 had 79% (RR: 0.21, 95% CI: 0.06, 0.75) and 86% (RR: 0.14, 95% CI: 0.04, 0.44) lower rates, respectively. We observed significantly lower rates in drop-in/respite program (RR: 0.23, 95% CI: 0.07, 0.80) and encampment sites (RR: 0.05, 95% CI: 0.004, 0.62) compared to shelter. The prevalence rate at encampments was significantly lower (RR: 0.04, 95% CI: 0.002, 0.63) compared to sites with a single room layout. Refugee sites showed a higher prevalence of SARS-CoV-2 compared to non-refugee sites, but the association was not significant (RR: 3.15, 95% CI: 0.68, 14.6)). The number of sporadic cases of COVID-19 in the community where the site was located was not associated with site confirmed positive cases.

**Table 2:** Association between each site characteristics and the number of daily confirmed COVID-19 cases (n=69)

|  | **Rate ratios (RR)** | **95 % CI** | **p-value** |
| --- | --- | --- | --- |
| Time period |  |  |  |
| *T1* | 1.00 | -- | -- |
| *T2* | 0.21 | 0.06, 0.75 | 0.016 |
| *T3* | 0.14 | 0.04, 0.44 | 0.001 |
| Shelter sector |  |  |  |
| Shelter | 1.00 | -- | -- |
| Drop-in/respite program | 0.23 | 0.07, 0.80 | 0.021 |
| Encampment | 0.05 | 0.004, 0.62 | 0.020 |
| COVID physical distancing sites | 1.12 | 0.34, 3.71 | 0.850 |
| Shelter layout |  |  |  |
| Single room | 1.00 | -- | -- |
| Shared room | 0.64 | 0.17, 2.48 | 0.518 |
| Open layout or dorm style | 0.66 | 0.17, 2.51 | 0.540 |
| Encampment (outdoor) | 0.04 | 0.002, 0.63 | 0.022 |
| Shelter group by age |  |  |  |
| Families/youth | 1.00 | -- | -- |
| Single adult | 0.44 | 0.13, 1.50 | 0.19 |
| Shelter group by gender |  |  |  |
| Men | 1.0 | -- | -- |
| Women | 1.21 | 0.32, 4.67 | 0.779 |
| Mixed | 0.99 | 0.36, 2.71 | 0.980 |
| Community transmission<sup>a</sup> | 0.97 | 0.90, 1.04 | 0.365 |
| Refugee site | 3.15 | 0.68, 14.6 | 0.143 |

Rate ratios and 95% confidence intervals estimated by fitting univariate negative binomial models including the site characteristics and log of total number of tests as the offset
<sup>a</sup> For every increase of 10 community cases

**Figure 1:**
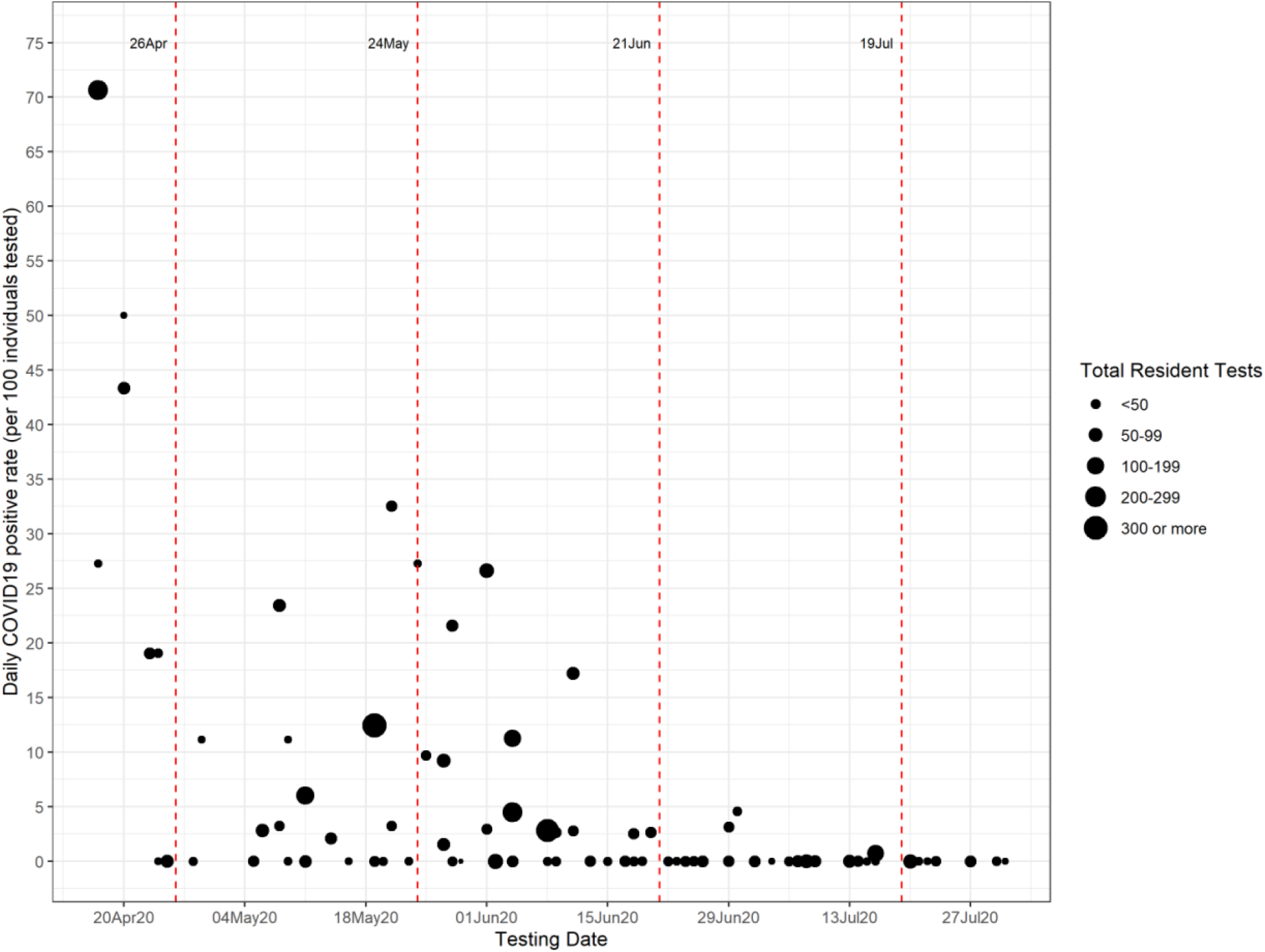
Daily positivity rates from April 1, to July 31, 2020. The size of the circles represents the number of residents tested on each day

**Figure 2:**
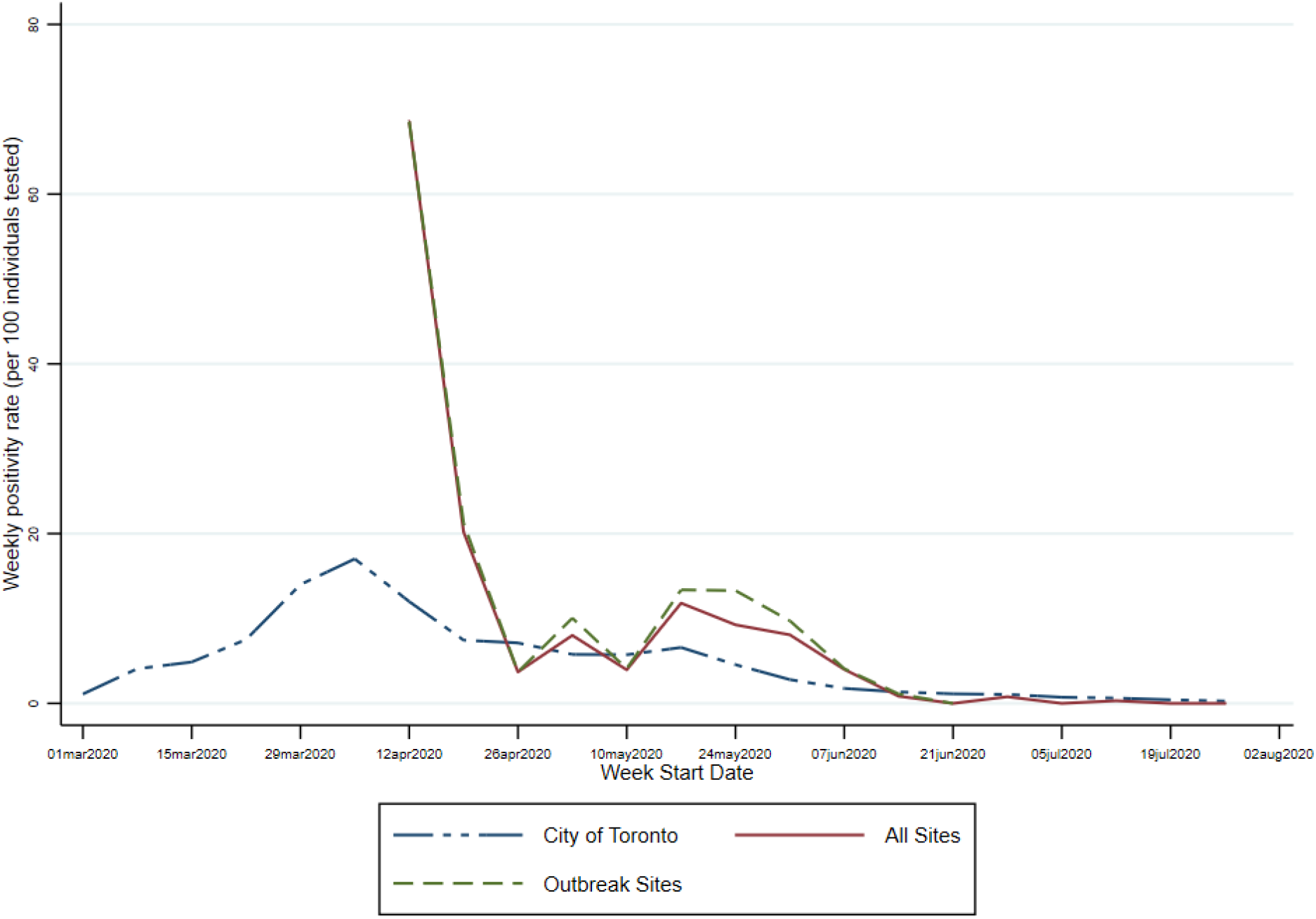
Weekly COVID-19 positivity rates (per 100 individuals tested) for the weeks starting on March 1, 2020 and ending on the week starting on July 26, 2020

### Interpretation

The study found great heterogeneity (0% to 70.4%) in the prevalence of SARS-CoV-2 infection across shelters in Toronto during the period April 17 to July 31, 2020. We found significantly higher positivity rates during the T1 (April 17-April 25) compared to later testing periods. We also found that site-based active case finding (testing in the absence of an outbreak) identified few cases and likely provided minimal benefits in terms of reducing the transmission of SARS-CoV-2. This finding is consistent with modeling studies that indicated that mass testing has to occur every 7 days to be a useful part of shelter COVID management (Baggett, Scott, et al., 2020). Positivity rates were significantly lower at drop-in/respite programs and encampment sites than at shelter sites.

This study provides insights into SARS-CoV-2 infection rates and shelter-level factors associated with positivity rates in the homeless population in Toronto. Our prevalence (8.5%) is higher compared to previous Ontario-based studies that used administrative health records to ascertain positivity rates (2.3 to 6.4%) in homeless populations (Richard et al., 2021; Wang et al., 2020). Since our study sample was drawn from sites serving people experiencing homelessness rather than users of the healthcare system who were homeless, our estimate may better reflect the positivity rate among people experiencing homelessness. Our rate is lower compared to prior studies conducted in shelters experiencing outbreaks, which found rates ranging from 18.0% to 41.7% (Baggett, Keyes, et al., 2020; Redditt et al., 2020; Tobolowsky et al., 2020). In contrast, we included active case finding and outbreak testing, which likely contributed to the observation of lower positivity rates.

The rapid decline in positivity rates over time may be attributed to the efficient coordination between shelter providers, TPH and hospital partners to screen and isolate infected residents at a COVID-19 recovery site and likely thus reducing the transmission of SARS-CoV-2. There is growing evidence that poor ventilation in enclosed indoor spaces may increase risk of COVID-19 transmission (Bhagat et al., 2020), and this may explain why we observed lower positivity rates in encampments. However, given the small sample size of encampment residents, this finding should be interpreted with caution.

The study has limitations. We used aggregate data at the site level and thus, individual-level characteristics such as symptoms, age and other sociodemographic characteristics could not be examined. There is a possibility of sampling bias, as sites chosen for testing based on identification of positive cases or at the request of the shelter providers. In addition, at some sites, the proportion of eligible residents who agreed to be tested was low. Given our sample size, we did not control for potential confounders in our models.

## Conclusions

Our study found a high prevalence of SARS-CoV-2 infection among people experiencing homelessness in Toronto during the first wave of the COVID-19 pandemic. This highlights the urgent need to implement measures to end homelessness, including by creating more affordable and supportive housing, increasing social assistance rates, and providing ongoing health and social support to individuals and communities most impacted by COVID, as well as implementing interim measures to decrease COVID transmission risk such as providing sufficient shelter space to allow for physical distancing, secure employment and paid sick days for staff, and sufficient PPE. Future work should examine the effectiveness of relocation of residents to COVID-19 response hotels/sites and other safety measures to ensure appropriate physical distancing in shelters on COVID-19 transmission.

### Contribution to knowledge

- This study provides insights into SARS-CoV-2 positivity rates and the site characteristics associated with an increased in positivity rates in the homeless population in Toronto, Canada
- Our finding highlights the urgent need to implement timely and coordinated crisis management to prevent and mitigate the negative consequences of the COVID-19 epidemic in the homeless population and those who serve them. Long-term strategies need to be in place to protect the homeless community in the event of a similar crisis in the future by creating more affordable and supportive housing, providing sufficient shelter space to avoid overcrowding, offering social assistance supports and developing evidence-informed outbreak and infection control guidelines.

## Data Availability

Data are available from Ontario Health Toronto upon reasonable request.

## Acknowledgements

We thank Rick Wang for creating the study figures. We also thank the people involved in the shelter outreach testing for their hard work, which allowed for timely data collection. We also would like to acknowledge Leona Pereira from Ontario Health Toronto, who provided expertise and knowledge on shelter testing coordination.

